# Temporal Trend in Mortality from COVID-19 Associated with Cardiovascular Disease

**DOI:** 10.1101/2023.04.26.23289183

**Authors:** Alexandra Sarau, Shengyuan Luo, Elaine Yi-Shuan Chen, Susan Gawel, Pankaja Desai, Nagarjuna Tippireddy, Anil J. Saldanha, Tisha Suboc, Gavin Cloherty, Alan Landay, Annabelle Santos Volgman

## Abstract

**Background:** The COVID-19 pandemic has endured for over three years with over twelve variants afflicting humans worldwide. The impact and longevity of the pandemic has driven the medical community and researchers to identify high-risk populations, yet few studies have explored temporal trends during the pandemic. The objective of this study is to investigate trends in all-cause mortality associated with co-morbid cardiovascular disease (CVD) throughout the consecutive waves of the COVID-19 pandemic.

**Methods:** A retrospective cohort study was conducted on patients with COVID-19 infection who received care at a tertiary care center in Chicago between March 2020 and September 2022.

Multivariable logistic regression was used to investigate associations between co-existing CVD (defined as congestive heart failure, myocardial infarction, cerebrovascular disease, peripheral vascular disease) and mortality, adjusting for age, sex, race, and comorbidities defined in the Charlson comorbidity index.

**Results:** The study included 38651 participants (mean age 45 years, 57.6% female, 11.4% with co-existing CVD). All-cause mortality in COVID-19 patients was highest during the Delta wave and remained elevated until the late Omicron wave. Mortality associated with co-existing CVD increased during the early pandemic waves, decreased in the later waves, but remained elevated relative to the overall population. When adjusted for age, sex, and race, all-cause mortality was 2.3-fold higher in patients with co-existing CVD compared to those with non-CVD comorbidities (OR = 2.30, 95% CI 1.95 – 2.62; p < 0.001).

**Conclusions:** While overall mortality rates declined toward the later waves of the pandemic, all-cause mortality associated with CVD remained elevated compared to individuals without CVD.

**Clinical Perspective:** *What is new?:*

- This study found that individuals with co-existing CVD have 2.3-fold higher risk of all-cause mortality compared to those with non-CVD comorbidities.
- Mortality associated with co-existing CVD was highest during the early waves of the pandemic and declined during the later waves, yet remained elevated relative to the overall population.

*What are the clinical implications?:*

- Increased all-cause mortality in COVID-19 patients with co-morbid CVD may be multifactorial from socioeconomic, psychosocial, behavioral, and biological variables.
- Individuals with co-existing CVD who become infected with COVID-19 should be considered an at-risk population for increased mortality.

## Introduction

The impact of Coronavirus disease 2019 (COVID-19) has touched millions of lives worldwide, causing downstream effects at the level of healthcare systems, communities, and patients. The COVID-19 pandemic has persisted for over three years, from March 2020 until the present day, with over twelve SARS-CoV-2 variants afflicting the globe. Consequently, we have had the opportunity to learn about the long-term effects of COVID-19 infections, examine temporal trends, and identify high-risk populations.

Much of the research on COVID-19 mortality describes associations within a specific interval or wave of the pandemic. For example, retrospective studies and meta-analyses have repeatedly shown that advanced age and cardiovascular disease (CVD) are associated with worse overall morbidity and mortality outcomes in those infected with COVID-19 ^1-4^. However, few studies have described the trends in COVID-19 mortality over time, starting from the emergence of the pandemic to the present day. A recent analysis of age-standardized death rates from 2010 to 2020 found that heart disease death rates declined by 9.8% from 2010 to 2019; however, death rates increased by 4.1% in 2020, coinciding with the advent of the pandemic ^5^. Heart disease death rates in 2020 were even more pronounced among non-Hispanic Black adults, with an increase of 11.2%. Similarly, a cohort study by Sidney et al. noted an increase in risk-associated death from heart disease and stroke, with the highest risk observed in racial and ethnic minority groups, especially non-Hispanic Black and Hispanic individuals ^6^.

The etiology of this unfavorable trend is not well defined. Still, multiple theories include reduced access to preventive health care, psychosocial and economic factors, increased stress and anxiety, diminished physical activity, and a possible predisposition to CVD in those with prior COVID-19 infection.

This study specifically investigates all-cause mortality associated with CVD through the sequential waves of the pandemic. It is important that trends associated with specific comorbidities, especially CVD, are detected as it has implications from not only a healthcare provider perspective but also from a public health and healthcare systems outlook.

## Methods

### Study Design

We performed a retrospective study among a cohort of patients who were under investigation for COVID-19 at Rush University System for Health from March 2020 to September 2022. We included patients greater than 18 years of age with a documented infection of COVID-19. We divided the pandemic into six distinct waves based on the predominant coronavirus variants and COVID-19 vaccine distribution. The study was approved by the Rush University Institutional Review Board (IRB) and exempt from informed consent.

### Exposure

CVD was defined using the International Classification of Diseases (ICD)-10 codes: congestive heart failure (CHF), myocardial infarction (MI), cerebrovascular disease (CEVD), or peripheral vascular disease (PVD).

### Outcomes

The primary outcome of this study is all-cause mortality, including both inpatient and outpatient deaths, as documented through a retrospective review of patients’ electronic medical records. Inpatient deaths were defined as deaths associated with in-hospital admissions or emergency room visits; outpatient deaths were defined as deaths associated with out-of-hospital or patient vehicle encounters. The date of death was identified through a retrospective review of the patient record. The secondary outcome of this study is inpatient hospitalizations.

### Statistical Analysis

Patient characteristics were summarized using descriptive statistics. We assessed associations between co-existing CVD and mortality using multivariable logistic regression, adjusting for age, sex, race, CVD comorbidities defined in the Charlson comorbidity index (CHF, MI, CEVD, and PVD), and non-CVD comorbidities defined in the Charlson comorbidity index (dementia, chronic pulmonary disease, rheumatic disease, peptic ulcer disease, mild liver disease, diabetes without chronic complication, diabetes with chronic complication, hemiplegia or paraplegia, renal disease, any malignancy [including lymphoma and leukemia, except malignant neoplasm of skin], moderate or severe liver disease, metastatic solid tumor, and AIDS/HIV) ^7^. A p-value of p < 0.05 was used to determine statistical significance. Age-stratified analyses were conducted to identify differences in demographics and comorbidities across age groups (18-49 years, 50-69 years, and 70-90 years). All statistical analyses were performed using R4.1.2.

## Results

Among 38,651 patients (mean age 45 years, 57.6% female, 11.4% with existing CVD comorbidities), 1,046 (2.7%) died. Across all waves of the pandemic, higher mortality was seen among individuals aged 70-90 (11.7%) and aged >90 (24.5%), men (3.5%), and persons with co-existing CVD comorbidities (12.2%) (**Table 1**). All-cause mortality in patients with COVID-19 was highest during the Delta wave (3.8% from June 2021 to December 2021) (**Table 2**) and remained elevated until the late Omicron wave (until March 2022). Mortality associated with co-existing CVD increased similarly through the early pandemic waves. Although decreasing in later waves, the mortality for patients with co-existing CVD remained elevated relative to the overall adult population (**Figure 1**). When adjusted for age, sex, and race, all-cause mortality was 2.3-fold higher in patients with co-existing CVD compared to those without any comorbidities as defined by the Charlson comorbidity index (OR = 2.30, 95% CI 1.95 – 2.62; p < 0.001) (**Figure 2a**). Further analysis stratified by age groups (adults with age <50 years, age 50-69, and age 70-90) indicate that co-existing CVD had a markedly increased risk of death amongst those <50 years of age (OR=8.26, 95% CI 5.42 – 12.55) (**Figure 2b**).

**Table 1:** Baseline characteristics of COVID-19-positive patients treated at Rush University System for Health from March 2020 to September 2022.

| Characteristic | N (%)<br>N=38651 | Death<br>N (%)<br>N=1046 | No Death<br>N (%)<br>N=37605 |
| --- | --- | --- | --- |
| **Age (years)** |  |  |  |
| 18 - 49 | 23990 (62.1%) | 125 (0.5%) | 23865 (99.5%) |
| 50-69 | 10540 (27.3%) | 404 (3.8%) | 10136 (96.2%) |
| 70-90 | 3860 (10.0%) | 453 (11.7%) | 3407 (88.3%) |
| > 90 | 261 (0.7%) | 64 (24.5%) | 197 (75.5%) |
| **Gender** |  |  |  |
| Female | 22262 (57.6%) | 475 (2.1%) | 21787 (97.9%) |
| Male | 16389 (42.4%) | 571 (3.5%) | 15818 (96.5%) |
| **Race** |  |  |  |
| White | 13008 (33.7%) | 415 (3.2%) | 12593 (96.8%) |
| Black or African American | 11560 (29.9%) | 316 (2.7%) | 11244 (97.3%) |
| Asian or Other | 1167 (3.0%) | 32 (2.7%) | 1135 (97.3%) |
| Undisclosed | 12916 (33.4%) | 283 (2.2%) | 12633 (97.8%) |
| **Comorbidity\*** |  |  |  |
| CVD | 4391 (11.4%) | 535 (12.2%) | 3856 (87.8%) |
| Myocardial infarction | 1089 (2.8%) | 165 (15.2%) | 924 (84.8%) |
| Congestive heart failure | 2120 (5.5%) | 288 (13.6%) | 1832 (86.4%) |
| Peripheral vascular disease | 1593 (4.1%) | 187 (11.7%) | 1406 (88.3%) |
| Cerebrovascular disease | 1836 (4.8%) | 255 (13.9%) | 1581 (86.1%) |
| Non-CVD | 11784 (30.5%) | 789 (6.7%) | 10995 (93.3%) |
| Dementia | 560 (1.4%) | 99 (17.7%) | 461 (82.3%) |
| Chronic pulmonary disease | 4541 (11.7%) | 228 (5.0%) | 4313 (95%) |
| Rheumatic disease | 606 (1.6%) | 38 (6.3%) | 568 (93.7%) |
| Peptic ulcer disease | 250 (0.6%) | 32 (12.8%) | 218 (87.2%) |
| Mild liver disease | 1434 (3.7%) | 99 (6.9%) | 1335 (93.1%) |
| Diabetes without chronic complication | 3502 (9.1%) | 208 (5.9%) | 3294 (94.1%) |
| Diabetes with chronic complication | 2089 (5.4%) | 239 (11.4%) | 1850 (88.6%) |
| Hemiplegia or paraplegia | 202 (0.5%) | 23 (11.4%) | 179 (88.6%) |
| Renal disease | 2253 (5.8%) | 319 (14.2%) | 1934 (85.8%) |
| Malignancy† | 1825 (4.7%) | 152 (8.3%) | 1673 (91.7%) |
| Moderate or severe liver disease | 199 (0.5%) | 45 (22.6%) | 154 (77.4%) |
| Metastatic solid tumor | 505 (1.3%) | 115 (22.8%) | 390 (77.2%) |
| AIDS/HIV | 142 (0.4%) | 7 (4.9%) | 135 (95.1%) |

\*CVD and non-CVD comorbidities are not mutually exclusive.
†Malignancy includes lymphoma and leukemia, except malignant neoplasm of skin.
Abbreviations: acquired immunodeficiency syndrome, AIDS; cardiovascular disease, CVD; human immunodeficiency virus, HIV.

**Table 2:** COVID-19 pandemic waves, hospitalization rates, and mortality rates of COVID-19 adult patients overall and with co-existing CVD

| Wave<br>Predominant<br>COVID-19 Variant | Overall Adult Population |  |  | Adults With Co-Existing CVD |  |  |
| --- | --- | --- | --- | --- | --- | --- |
|  | N | Hospitalization<br>N (%) | Mortality<br>N (%) | N | Hospitalization<br>N (%) | Mortality<br>N (%) |
| 1:<br>Wildtype+D614G<br>(3/7/2020 –<br>3/20/2021) | 21253 | 5901; 27.8% | 627; 3.0% | 2130 | 1142; 53.6% | 297;<br>13.9% |
| 2: Alpha<br>(3/21/2021 –<br>6/19/2021) | 1871 | 801; 42.8% | 57; 3.0% | 247 | 168; 68.0% | 38; 15.4% |
| 3: Delta<br>(6/20/2021 –<br>12/11/2021) | 3070 | 1259; 41.0% | 118; 3.8% | 372 | 229; 61.6% | 56; 15.1% |
| 4: Omicron BA.1<br>(12/12/2021 –<br>3/19/2022) | 8236 | 2290; 27.8% | 204; 2.5% | 1028 | 552; 53.7% | 119;<br>11.6% |
| 5: Omicron BA.2<br>(3/20/2022 –<br>6/18/2022) | 1750 | 469; 26.8% | 18; 1.0% | 232 | 119; 51.3% | 12; 5.2% |
| 6: Omicron BA.4/<br>BA.5<br>(6/19/2022 –<br>(9/30/2022) | 2471 | 797; 32.3% | 22; 0.9% | 382 | 184; 48.2% | 13; 3.4% |
| Overall | 38651 | 11517; 29.8% | 1046;<br>2.7% | 4391 | 2394; 54.5% | 535;<br>12.2% |

**Figure 1:**
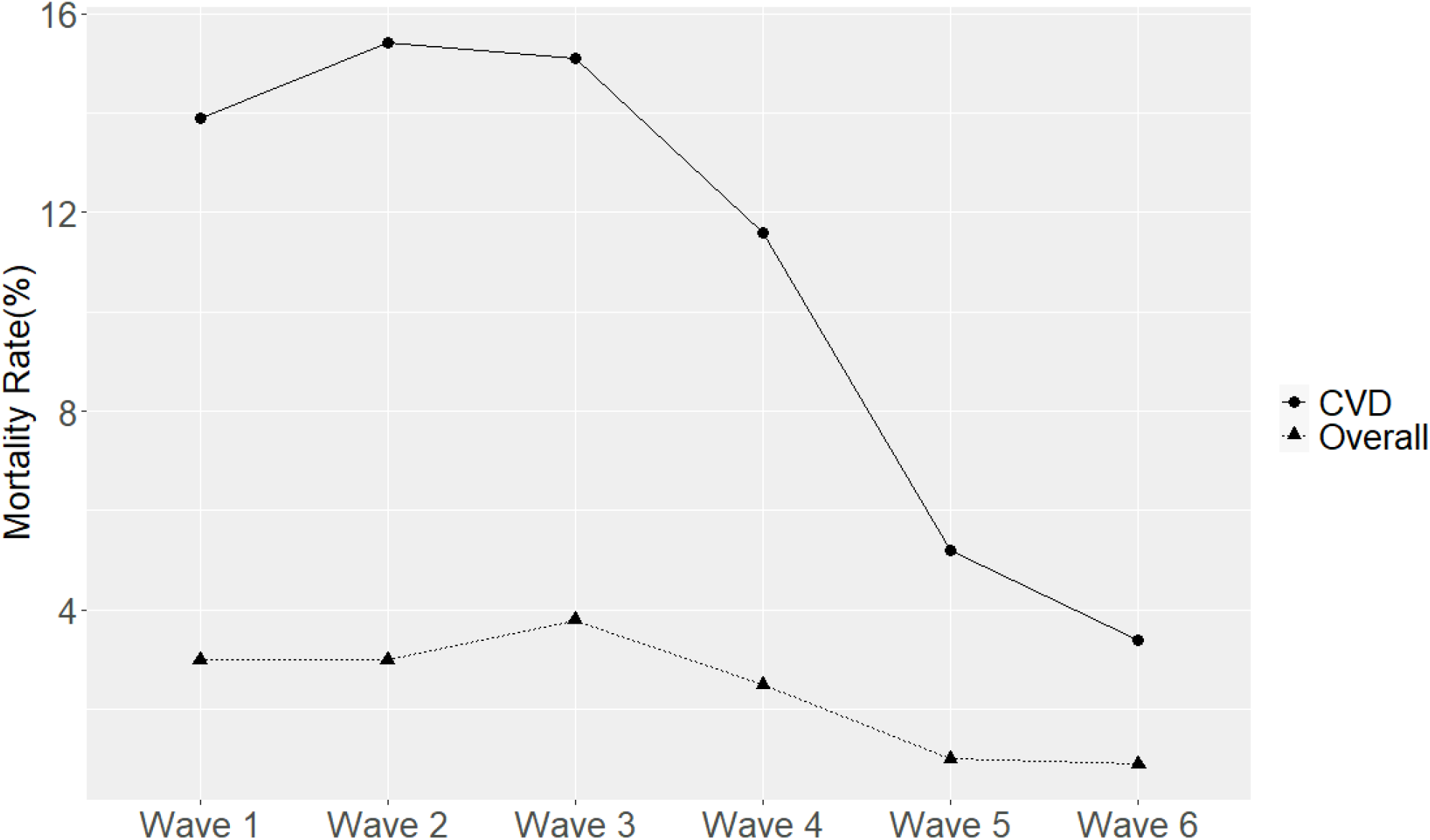
Mortality rate in each pandemic wave (Overall vs. co-existing cardiovascular disease)

**Figure 2:**
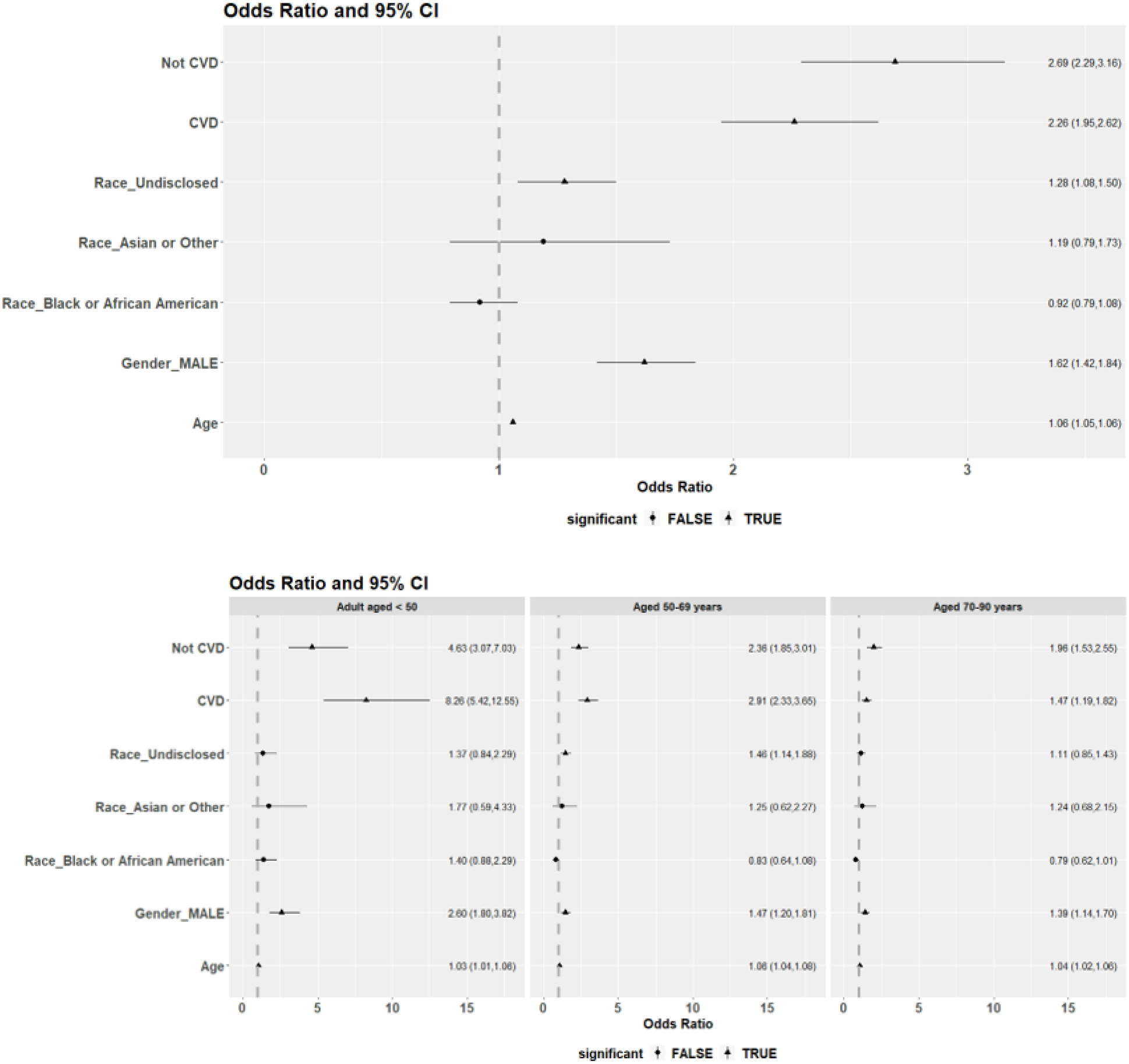
Multivariable logistic regression model for **(a)** overall mortality and **(b)** age-stratified mortality.

Further analyses were performed to evaluate the four CVD comorbidities in the Charlson index (CHF, MI, CEVD, and PVD). Mortality in patients with COVID-19 was highest during the Alpha wave for CEVD (18.4%), CHF (19.8%), and PVD (15.8%). For those with a history of MI, the highest mortality occurred during the initial Wildtype variant (18.2%) (**Figure 3, Table 3**), plateaued through Wave 2 – 4 but began rising again during Wave 6. In contrast, mortality in patients with co-existing CEVD, CHF, or PVD continued to decline over time. Hospitalizations associated with co-existing CVD similarly peaked with the Alpha wave and generally declined with subsequent waves (**Figure 3, Table 3**).

**Figure 3:**
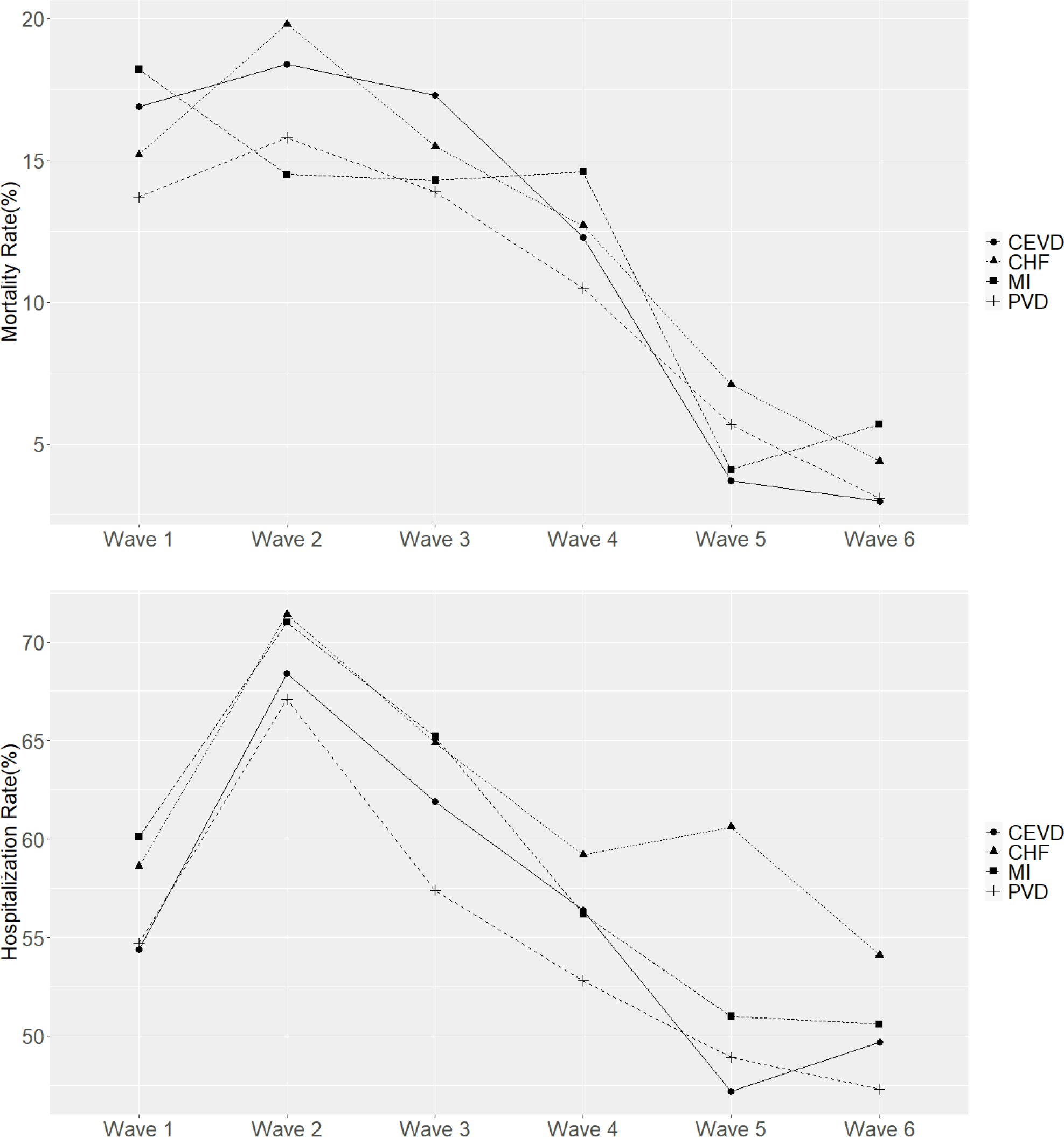
Mortality (top) and hospitalization (bottom) rate in each pandemic wave (Overall vs co-existing CVD) Abbreviations: cerebrovascular disease, CEVD; congestive heart failure, CHF; myocardial infarction, MI; peripheral vascular disease, PVD.

**Table 3:**
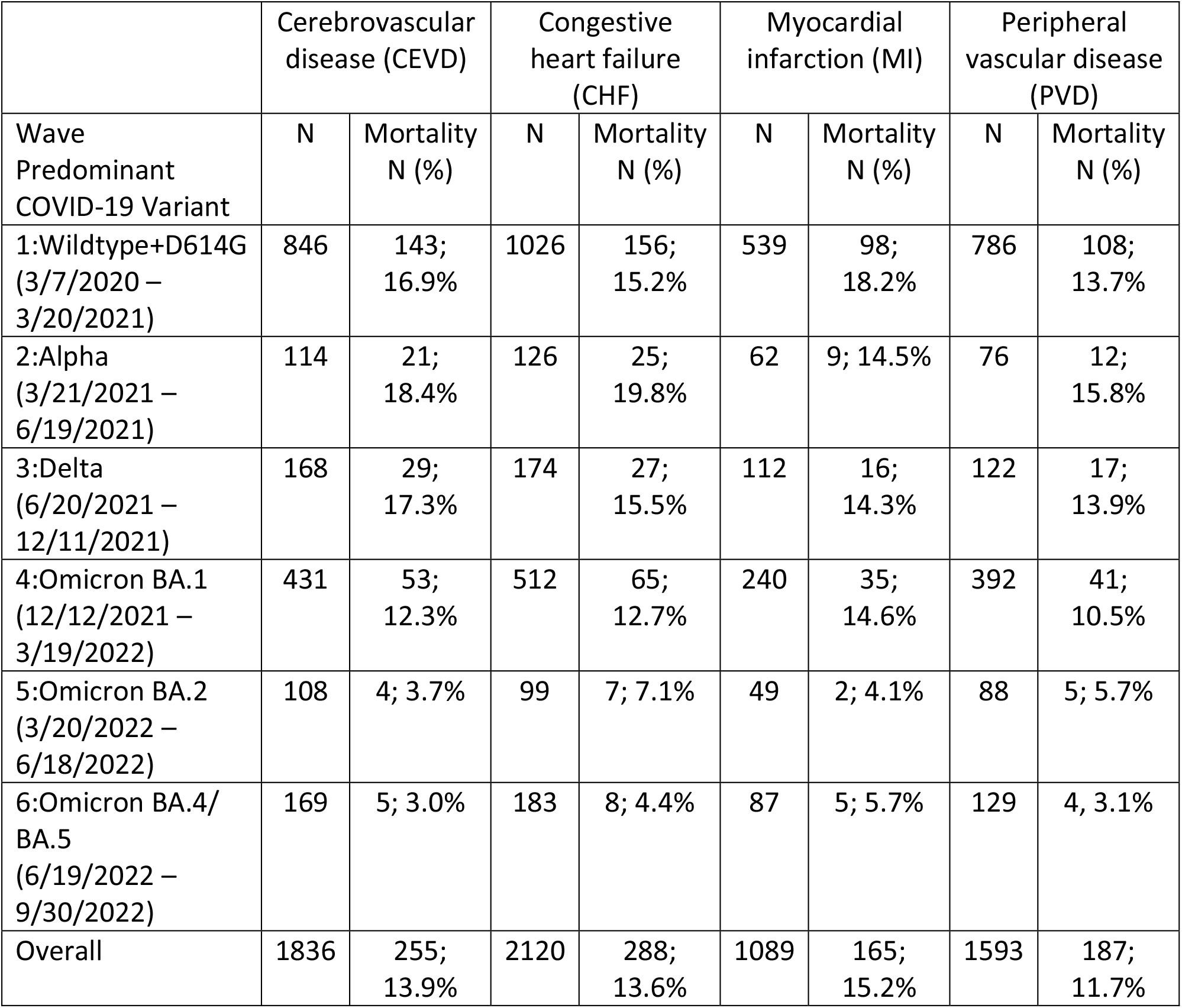
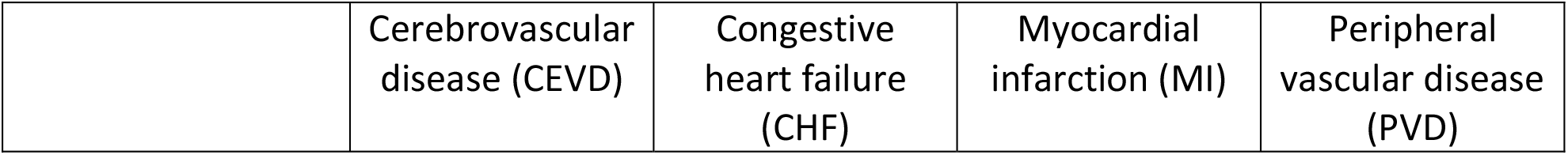
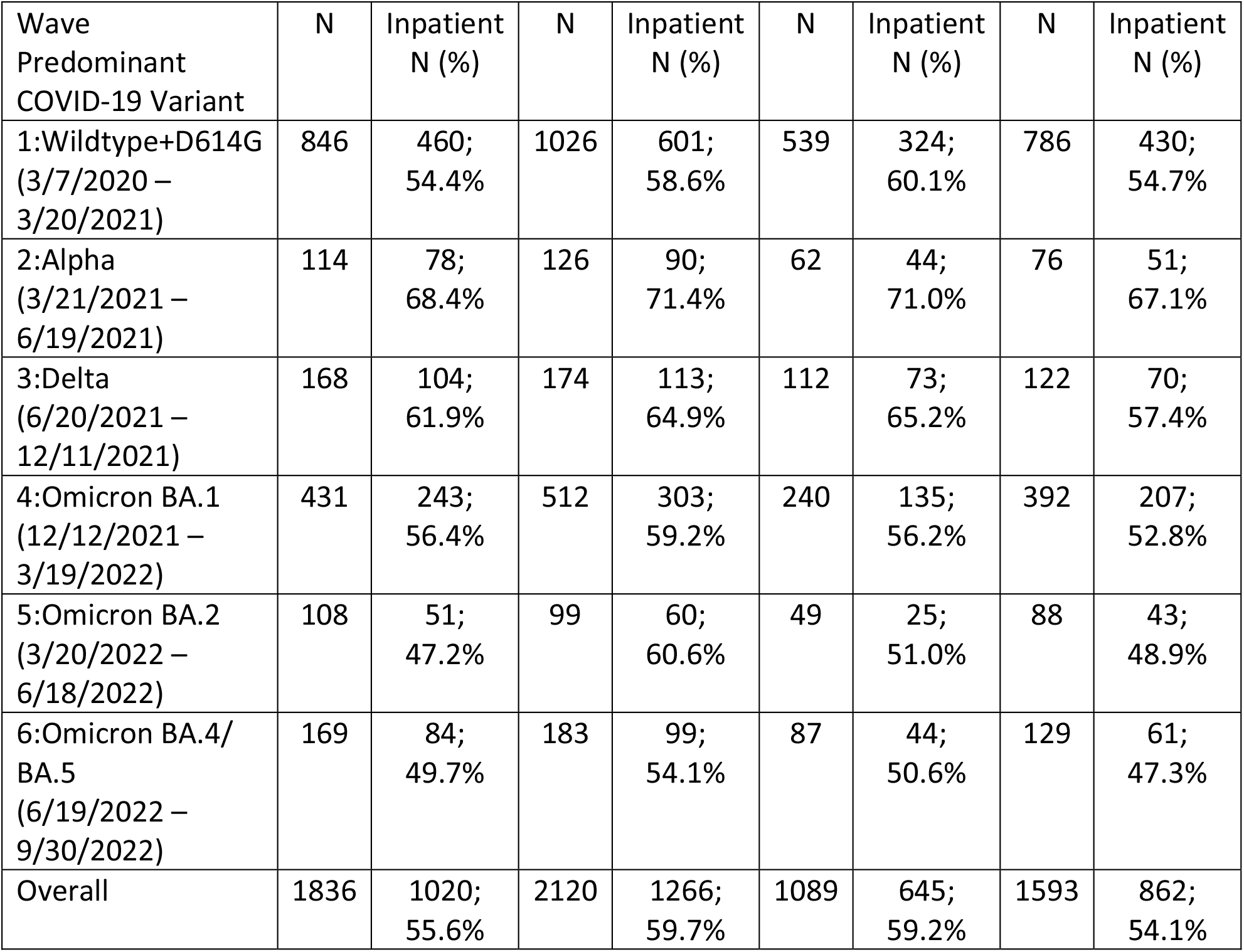
COVID-19 pandemic waves, mortality (top), and hospitalization (bottom) rates of COVID-19 adult patients with co-existing CVD by CVD type.

|  | Cerebrovascular disease (CEVD) |  | Congestive heart failure (CHF) |  | Myocardial infarction (MI) |  | Peripheral vascular disease (PVD) |  |
| --- | --- | --- | --- | --- | --- | --- | --- | --- |
| Wave<br>Predominant<br>COVID-19 Variant | N | Mortality<br>N (%) | N | Mortality<br>N (%) | N | Mortality<br>N (%) | N | Mortality<br>N (%) |
| 1:Wildtype+D614G<br>(3/7/2020 – 3/20/2021) | 846 | 143;<br>16.9% | 1026 | 156;<br>15.2% | 539 | 98;<br>18.2% | 786 | 108;<br>13.7% |
| 2:Alpha<br>(3/21/2021 – 6/19/2021) | 114 | 21;<br>18.4% | 126 | 25;<br>19.8% | 62 | 9; 14.5% | 76 | 12;<br>15.8% |
| 3:Delta<br>(6/20/2021 – 12/11/2021) | 168 | 29;<br>17.3% | 174 | 27;<br>15.5% | 112 | 16;<br>14.3% | 122 | 17;<br>13.9% |
| 4:Omicron BA.1<br>(12/12/2021 – 3/19/2022) | 431 | 53;<br>12.3% | 512 | 65;<br>12.7% | 240 | 35;<br>14.6% | 392 | 41;<br>10.5% |
| 5:Omicron BA.2<br>(3/20/2022 – 6/18/2022) | 108 | 4; 3.7% | 99 | 7; 7.1% | 49 | 2; 4.1% | 88 | 5; 5.7% |
| 6:Omicron BA.4/<br>BA.5<br>(6/19/2022 – 9/30/2022) | 169 | 5; 3.0% | 183 | 8; 4.4% | 87 | 5; 5.7% | 129 | 4, 3.1% |
| Overall | 1836 | 255;<br>13.9% | 2120 | 288;<br>13.6% | 1089 | 165;<br>15.2% | 1593 | 187;<br>11.7% |

| Wave<br>Predominant<br>COVID-19 Variant | N | Inpatient<br>N (%) | N | Inpatient<br>N (%) | N | Inpatient<br>N (%) | N | Inpatient<br>N (%) |
| --- | --- | --- | --- | --- | --- | --- | --- | --- |
| 1:Wildtype+D614G<br>(3/7/2020 –<br>3/20/2021) | 846 | 460;<br>54.4% | 1026 | 601;<br>58.6% | 539 | 324;<br>60.1% | 786 | 430;<br>54.7% |
| 2:Alpha<br>(3/21/2021 –<br>6/19/2021) | 114 | 78;<br>68.4% | 126 | 90;<br>71.4% | 62 | 44;<br>71.0% | 76 | 51;<br>67.1% |
| 3:Delta<br>(6/20/2021 –<br>12/11/2021) | 168 | 104;<br>61.9% | 174 | 113;<br>64.9% | 112 | 73;<br>65.2% | 122 | 70;<br>57.4% |
| 4:Omicron BA.1<br>(12/12/2021 –<br>3/19/2022) | 431 | 243;<br>56.4% | 512 | 303;<br>59.2% | 240 | 135;<br>56.2% | 392 | 207;<br>52.8% |
| 5:Omicron BA.2<br>(3/20/2022 –<br>6/18/2022) | 108 | 51;<br>47.2% | 99 | 60;<br>60.6% | 49 | 25;<br>51.0% | 88 | 43;<br>48.9% |
| 6:Omicron BA.4/<br>BA.5<br>(6/19/2022 –<br>9/30/2022) | 169 | 84;<br>49.7% | 183 | 99;<br>54.1% | 87 | 44;<br>50.6% | 129 | 61;<br>47.3% |
| Overall | 1836 | 1020;<br>55.6% | 2120 | 1266;<br>59.7% | 1089 | 645;<br>59.2% | 1593 | 862;<br>54.1% |

## Limitations

A limitation of this study is that this database could not determine if the mortality was due to the preexisting CVD in the patient or if it was due to the COVID-19 infection. Another limitation is that we used the date range to represent the COVID-19 variant, not the patients’ actual viral infection.

## Discussion

This retrospective cohort study highlights temporal trends in CVD-associated mortality compared to the general population of COVID-19 patients. While overall mortality rates declined toward the more recent waves of the pandemic with late Omicron variants predominating, all-cause mortality associated with CVD remained elevated compared to non-CVD-associated individuals. To our knowledge, this is the first study to report these findings and one of few studies that describe temporal trends during the pandemic. While a few papers illustrate CV mortality trends over the last decade, they did not specifically analyze patients with documented COVID-19 infections ^5,6^. A study performed in Japan did look at temporal trends related to in-hospital COVID-19 mortality during three waves of the pandemic and noted a progressive decline in in-hospital mortality with each wave when adjusted for sex and age; however, they did not look at CVD associations ^8^.

It is well known that cardiovascular comorbidities are associated with increased morbidity and mortality due to COVID-19 ^9^. However, whether individuals with prior COVID-19 infection are at increased risk of subsequent CVD events is still unclear. It has been suggested that patients with a prior COVID-19 infection are at increased risk of atrial fibrillation, venous thromboembolism, heart failure, and death; these events were noted particularly in patients who required hospitalization and more commonly in the early post-infection period within 30 days ^10^. Similar findings were observed in a study by Wang et al., who noted a significantly higher 12-month risk of CVD in survivors of COVID-19 compared to non-infected controls ^11^.

There are no proven explanations for the increase in all-cause mortality in COVID-19 patients with associated CVD; however, the etiology is likely multifactorial. Probable factors include lack of access to or reluctance to seek healthcare during the pandemic, healthcare disparities in certain racial/ethnic and socioeconomic groups, hospital strain and limited healthcare resources, reduced physical activity, increased propensity for detrimental habits, and a potential predisposition to CVD in those with prior COVID-19 infection. The variations of mortality rates in patients with a history of MI suggest that the variants of COVID-19 may have different virulence in atherosclerotic disease.

During the early waves of the pandemic, there was limited access to preventive health care and a shift toward telemedicine, which led to delays in detecting and treating cardiovascular conditions. A web-based survey of U.S. adults obtained in June 2020 revealed that 40.9% of adults avoided medical care due to COVID-19 concerns, with 12.0% avoiding urgent/emergent care and 31.5% avoiding routine care ^12^.

A recent study by Gotanda et al. describes a substantial decline in blood pressure monitoring during the early pandemic; they also found that blood pressure measurements in patients with chronic hypertension were significantly higher during the pandemic compared to pre-pandemic times ^13^. Furthermore, there was a delay in care from the perspective of hospital systems, including deferral of elective procedures, such as outpatient coronary angiograms and percutaneous coronary interventions. There was even an estimated 38.0% reduction in STEMI activations during the early waves of the pandemic ^14^. These factors likely propagated detrimental downstream cardiovascular sequelae such as congestive heart failure, MI, and cardiovascular mortality.

Multiple psychosocial and behavioral factors likely contributed to the temporal rise in CVD-associated mortality observed during the pandemic. Quarantine periods and social distancing became essential practices to mitigate the spread of COVID-19 infection; however, a recent scoping review revealed that social isolation and loneliness are directly associated with increased mortality from coronary heart disease and stroke ^15^. Due to lockdown and quarantine circumstances, many people were required to stay in their homes. Over time, the work culture for many occupations has transitioned to “work from home” conditions.

As a result, less participation in physical activity, increased alcohol consumption, higher anxiety scores, increased weight gain, and higher rates of obesity have been observed ^16,17^.

Further research is needed to explore the possible underlying mechanism(s) behind observed increases in cardiovascular conditions in patients with COVID-19. It would also be valuable to explore direct causes for the temporal rise in CVD-associated mortality, as well as identify other temporal trends in the COVID-19 population, as these would have significant public health implications.

## Data Availability

The data referred to in the manuscript is included in the tables and figures. Any raw form of the data can be included by request.

